# Coping strategies employed by transgender youth with higher and lower quality of life

**DOI:** 10.1101/2022.08.17.22278903

**Authors:** Ítala Raymundo Chinazzo, Anna Martha Vaitses Fontanari, Bruno de Brito Silva, Lucca P. Rodrigues, Angelo Brandelli Costa, Maria Inês Rodrigues Lobato

## Abstract

Transgender youth are especially susceptible and vulnerable to mental health concerns. Minority Stress Theory may explain these mental health concerns and avoidance to seek healthcare provision in this population. Understanding transgender youth adaptive and maladaptive coping mechanisms may help create strategies that promote quality of life and reduce the negative influence of stress on their mental health. Our study aims to measure the quality of life of Brazilian binary and non-binary transgender youth and analyze the association with their coping strategies used for dealing with general social and gender-related stress. Participants were recruited through Facebook advertisement directed to young Brazilians, aged from 16 to 24 years, who “liked” pages or joined groups related to LGBTQ+ movement. Gender identity was evaluated by the Two Steps Method. Coping was evaluated by the Coping with Stress Checklist; and quality of life, by the Quality of My Life questionnaire. The final sample consisted of 185 participants. Out of the total, 95 (46.34%) identified as transgender boys, 45 (21.95%) as transgender girls, and 65 (31.71%) as non-binary youth. The mean age was 18.61 years (SD 2.43). The study found that 53.9% of the sample had low self-perceived quality of life, 48.0% depression, and 68.3% anxiety. Better quality of life was related to socialization (p=0.02), whereas a worse quality of life was related to avoidance attitudes (p=0.05), concerning coping with general stress. The study found no association between coping strategies for gender-related stress and quality of life. The Brazilian transgender youth may need support to develop adaptive coping skills to deal with both general and gender-related stress. Also, social interventions against prejudice are needed to achieve better results in the quality of life for transgender youth. Mental health professionals should explore the unique needs and stressors of transgender identity and explore adaptive coping strategies.

## Introduction

The term transgender denotes individuals whose gender identity does not align with their sex assigned at birth, whereas the expression “cisgender” refers to individuals whose gender identity matches their sex assigned at birth (Giblon & Bauer, 2017). Transgender is an umbrella term that covers a broad continuum of gender identities, including binary and non-binary people. While binary people identify either as a woman or as a man – regardless of whether they are transgender or not – non-binary people recognize themselves as belonging to a third different gender, having more than one gender, or as being agender (Bass et al., 2018). Considering that non-binary people may seek gender-affirming care and that transgender people do not necessarily identify within the gender binaries, there are numerous intersections between non-binary and transgender. Nevertheless, since not all non-binary people identify as transgender, and vice versa, using both expressions is necessary (Bass et al., 2018; Coleman et al., 2012; Richards et al., 2016).

Transgender adults face high rates of violence throughout Brazil and the world (Balzer, Lagata, & Berredo, 2016; Benevides & Nogueira, 2019; Silva, 2018; Chinazzo et al. 2020), which not only directly affects their physical and mental health (Magno, Dourado, & Silva, 2018; Zucchi, 2019), but also limits their access to health services (Rocon et al. 2016; Rodriguez, 2014; Sousa & Iriart, 2018) and education (Associação Brasileira de Lésbicas, Gays, Bissexuais, Transexuais e Travestis [ABGLT], 2016). For instance, longitudinal studies suggest that transgender people have higher rates of completed suicide than their cisgender peers (Asscheman et al., 2011; Dhejne et al., 2011), and also experience a lower quality of life (Newfield et al.2006; Jones et al., 2019).

Transgender binary and non-binary youth are especially susceptible and vulnerable to mental health concerns (Chinazzo et al., 2020; Connolly et al., 2016; Dhejne et al., 2016; Perez-Brumer et al., 2015). Transgender youth from the United States are between 3.2 and 3.6 times more likely to have suicidal ideation and suicidal attempts, respectively, when compared to the general population (Reisner et al., 2015). Furthermore, according to their parents or caregivers, Canadian transgender youth talk 5.1 times more about suicide than their siblings (Aitken et al., 2016). Regarding non-binary transgender youth, Thorne et al. (2019) found evidence of a difference in their mental health (anxiety and depression) when compared with binary transgender youth, whereas Rimes et al. (2019) found no apparent differences when compared with binary transgender youth.

Mental health problems are associated with low quality of life in transgender people (Bouman et al., 2016). Quality of life is defined and related to an “individual’s perception of their position in life in the context of the culture and value systems in which they live and in relation to their goals, expectations, standards and concerns” (World Health Organization [WHO], 2021). Studies (Newfield et al., 2006; Nobili, Glazebrook & Arcelus, 2018) have reported that young transgender individuals have a lower perceived quality of life compared to the general population. Moreover, transgender youth have been associated with greater mental health problems (Arcelus et al, 2016; Heylens et al., 2014) and a lower mental health-related quality of life (Newfield et al., 2006). Additionally, the detrimental effects of body dissatisfaction in cisgender groups are observed as a factor for an increasing risk of depression (Paxton et al., 2006), low self-esteem (Davison & McCabe, 2006), and eating disorders (Stice & Shaw, 2002). Reports suggest that body dissatisfaction may be higher among transgender people (Witcomb et al., 2015; Ålgars, Santilla, & Sandnabba, 2010).

The Minority Stress Theory helps understanding these health disparities (Hendricks & Testa, 2012; Meyer, 1995, 2003). In addition to the general stress of life, belonging to a minority group increases chronic stress by enacted stigma, expectations of rejection, and internalization of social stigma, resulting in psychological distress and avoidance to seek healthcare provision. However, identification with a minority group may give access to community resources such as support groups, community centers, minority role models, as well as affirmative actions and specialized clinics (Meyer, 2015). Such resources are conceptualized as *minority coping*, which can buffer the effect of minority-specific stressors on the mental health of transgender individuals (DiFulvio, 2015; Stanton, Ali & Chaudhuri, 2017; Zucchi et al., 2019). Specifically, social support from one’s community – a resource derived from minority coping – seems to positively affect psychological well-being (Stanton, Ali, & Chaudhuri, 2017; Zucchi et al., 2019) as well as lessen the consequences of stigma on mental health (DiFulvio, 2015).

Stress is the reaction to a situation in which a person’s physical or psychological resources are challenged as aforementioned. Coping consists of the efforts that a person makes to adapt to stress and to manage external and/or internal demands that exceed the resources of the person. However, being able to cope does not necessarily translate as successfully adjusting. A person’s reactions in stressful environments are dependent on the person’s appraisal of a given situation, combined with the physical or psychological resources available (Lazarus & Folkman, 1984). The evaluation of this mechanism is adaptive or non-adaptative, depending on the ability to buffer or not the stresses they experience. Coping strategies can be learned, so it is important knowing which mechanisms are adaptive and, therefore, protective factors for mental health in transgender youth.

A protective factor for transgender individuals is the available social support (DiFulvio, 2015; Singh, Hays, & Watson, 2011; Puckett et al., 2019; Stanton, Ali, & Chaudhuri, 2017; Zucchi et al., 2019). This may be understood as a coping strategy for patients who have positive social relations, termed as “minority coping.” Social support means the existence of meaningful relations with people, in which there is trust, affection, and acceptance when sharing one’s life experience and feelings. There are two types of social support—informal and formal. The source of informal social support is the daily social network such as family members, friends, and social groups who help in everyday activities; whereas formal social support is related to social assistance organizations, professionals, and health services.

### Current Study

Most studies investigating coping strategies have focused on gay, lesbian, and transgender adults (Budge, Adelson, & Howard, 2013; Hughto, Pachankis, & Willie, 2017; Lehavot, 2012). Therefore, it is essential to evaluate transgender youth. Better understating of transgender youth coping mechanisms may help create strategies to promote well-being and reduce the negative influence of stress on their mental health. Our study aims to describe the coping behaviors employed by transgender youth who have a higher quality of life in comparison with those with a lower quality of life.

## Methods

### Study Sample

From February to April 2018, participants were recruited via Facebook advertisements directed towards Brazilian adolescents and young adults, aged 16–24, who “liked” pages or joined groups related to the LGBTQ+ movement. Gender identity was evaluated by the Two Steps Method (Bauer et al., 2017; Sausa et al., 2009). Firstly, sex at birth was assessed by the question: “What sex were you assigned at birth on your original birth certificate?”. Participants could choose between the “female” and “male” alternatives. Secondly, gender identity was evaluated as an open question “How do you currently describe your gender?”. All participants responded within the “woman,” “man,” and “non-binary” alternatives.

In total, 706 young people started the online protocol. Nevertheless, to guarantee the quality of the data, participants who did not reply to the two-step questions as transgender and non-binary were excluded. Likewise, duplicates and discrepant responses were removed. A total of 165 individuals were excluded because they did not complete the Coping with Stress Checklist. The final sample was composed of 185 participants in the data analysis. Fig 1 summarizes the participants’ flow.

**Fig 1.**
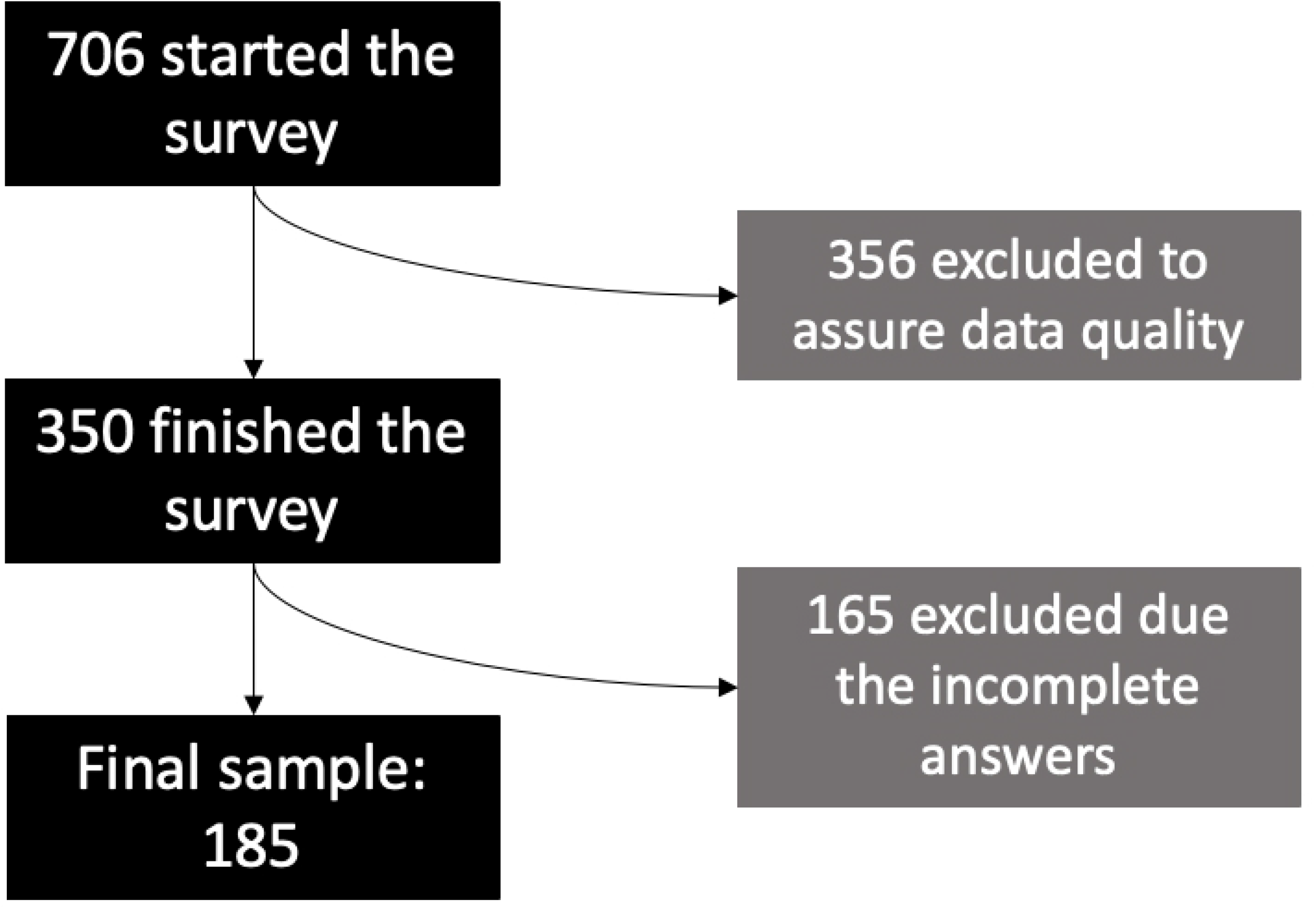
Participants’ sampling flowchart.

### Measures

The survey was inspired by the TransYouth CAN! Project (see http://transyouthcan.ca/). The cross-cultural adaptation for the Brazilian populations was based on the work by Borsa, Damásio, and Bandeira (2012); it is better described in previous publications (Fontanari et al., 2019).

Socioeconomic Status (SES) was measured using the Deprivation Scale, created by the “TransYouth CAN! Team” to access the information about the basic needs of our sample. The Deprivation Scale consisted of five questions asking whether participants had access to school supplies, internet, proper seasonal clothing, other essential clothing, and trustworthy transportation in a one-year period. Each question could be responded using a five-point Likert scale ranging from “never” (one point) to “always” (five points), in which a higher score represents less deprivation. The score was calculated by the arithmetical mean of the five questions (mean = 4.31; SD = 0.72); the Cronbach’s alpha coefficient was 0.820.

Coping with Stress Checklist, developed by the TransYouth CAN! Team is a questionnaire that assumes that young transgender people deal with a two-tier “stress situation.” The first one is the stress that happens in everyday life, identified as General Stress; and the second one is identified as Gender-Related Stress linked to their transgender condition. The questionnaire is composed of a 26-item list containing adaptive and maladaptive coping strategies, which assess the coping style and how each person deals with the General Stress and Gender Related Stress in a 30-day period. The 26-items were divided by the research team into four dimensions for each type of stress: 1. hobby strategies (watching movies, TV or on-line videos, playing videogame, and reading), 2. socialization (talking with LGBT and non-LGBT friends, spending time with family, volunteer work, spending time in social events, and publishing about themselves on on-line social networks), 3. introspection (physical exercises, deep breathing/relaxation exercises, writing a diary, praying, meditation) and 4. avoidance (excessive sleeping, eating, shopping, cosmetic self-care, day dreaming, looking for noncommitted partners, and ignoring or pretending that the stress is not real). For each strategy, participants could choose between “yes” and “no” alternatives. Cronbach’s alpha value of general stress is 0.74, and gender-related stress is 0.88.

To assess quality of life, the first item of the Quality of My Life (QoML) Questionnaire states “overall, my life is…” and is answered with a visual analog scale (Bowling, 2005; Feldman et al., 2000). There are five response options on this scale: excellent, very good, good, fair, or poor. Responses were dichotomized into “poor” and “excellent” quality of life, using the median.

The Overall Anxiety Severity and Impairment Scale (OASIS) (Norman et al., 2006) is a brief continuous measure of five questions about the level of anxiety and prejudice that participants felt in a seven-day period. Each question was measured on a five-point Likert scale, ranging from 0 (no anxiety or impairment from anxiety) to 4 (constant anxiety or impairment from anxiety). The responses were dichotomized and the cut-off point was 8, in which participants who scored more than 8 were more likely to have an anxiety diagnosis.

The Modified Depression Scale (MDS) (Dunn, Johnson & Green, 2012) was used to assess the frequency of five depressive symptoms in a 30-day period: “in the last 30 days how often have you… (1) felt very sad, (2) felt very angry or in a bad mood, (3) felt hopeless about the future, (4) slept much more or much less than usual, and (5) had difficulty concentrating on your studies.” Each question was answered on a five-point Likert scale: 1 (never), 2 (rarely), 3 (sometimes), 4 (often), and 5 (always). The responses were dichotomized according to the cut-off point of 17, in which participants who scored more than 17 were more likely to have a diagnosis of depression.

To evaluate gender dysphoria, a version of the Utrecht Gender Dysphoria Scale was used – adapted by the Trans Youth Can! Team, in which some items were added or deleted, making it more appropriate for non-binary persons. The scale consisted of 15 items, covering dysphoria related to the body (e.g., I wish I had been born in a different body) and social life (e.g., it bothers me to be called by the wrong gender). The items were answered on a five-point Likert scale, ranging from “totally disagree” to “totally agree,” plus the answer option “does not apply to me.” The score was obtained by summing up all items; for analysis the answers were divided into whether the participants identified as being without or almost without gender dysphoria, or with gender dysphoria or with a lot of gender dysphoria.

### Statistical Analysis

The software SPSS 18.0 was used to perform statistical analysis. Firstly, background characteristics were compared between those reporting in QoML, which was divided into poor and excellent life quality. Secondly, a Poisson Regression in two steps was applied to compare the four dimensions of coping strategy, from the Coping with Stress Checklist, with Quality of My Life, controlled by sociodemographic data. In the first step of the logistic regression, only the four dimensions of coping with stress checklist were included. In the second step, the variables age, SES, anxiety, depression, and gender dysphoria were also included. Both steps were performed for coping strategies with general stress and stress related to gender identity.

### Ethical Considerations

The Ethical Committee and Research Commission of the Federal University of Rio Grande do Sul Psychology Institute approved the project (14221513.4.0000.5334). All participants signed an online ethical consent form; for participants aged under 18 years, a consent term was provided by their guardians.

## Results

For this study, 185 participants completed the Coping with Stress Checklist. Among them, 90 (48.6%) identified as transgender boys, 37 (20.0%) as transgender girls, and 58 (31.4%) as non-binary persons (Table 1). The mean age was 18.50 years (SD 2.44) and youth with an average quality of life did not differ in sociodemographic characteristics.

**Table 1.** Sample description.

|  |  | N | % |
| --- | --- | --- | --- |
| Gender identity | Male | 90 | 48.6 |
|  | Female | 37 | 20.0 |
|  | Non-binary | 58 | 31.4 |
| Education | School | 70 | 37.8 |
|  | College preparatory programs | 13 | 7.0 |
|  | Technical course | 13 | 7.0 |
|  | College | 50 | 27.0 |
|  | Not studying | 39 | 21.1 |
| State | São Paulo | 51 | 28.5 |
|  | Rio Grande do Sul | 37 | 20.7 |
|  | Rio de Janeiro | 19 | 10.6 |
|  | Minas Gerais | 15 | 8.4 |
|  | Other | 57 | 31.8 |
| **Time living in perceived gender** | All the time | 86 | 46.5 |
|  | Half the time | 68 | 36.8 |
|  | Never | 31 | 16.8 |
| **Legal identity transition** | Social card | 14 | 7.6 |
|  | Identity card | 2 | 1.1 |
|  | Social and identity card | 2 | 1.1 |
|  | It is in process | 88 | 47.6 |
|  | Does not plan to change name | 18 | 9.7 |
|  | Prefer not to answer | 61 | 33.0 |
| **Public knowledge of your transgender identity** | Always | 18 | 9.7 |
|  | Almost always | 32 | 17.3 |
|  | About half the time | 34 | 18.4 |
|  | Rarely | 58 | 31.4 |
|  | Never | 23 | 12.4 |
|  | I do not know | 19 | 10.3 |
| **Hormone use** | Yes, regularly | 35 | 19.0 |
|  | Yes, once or twice | 14 | 7.6 |
|  | Never | 125 | 67.9 |
|  | I do not know / I would rather not answer | 10 | 5.5 |
| **Hormone use over time** | I am not looking for hormonal treatment | 85 | 46.2 |
|  | < 1 year | 56 | 30.4 |
|  | 1 - 2 years | 22 | 12.0 |
|  | 2 - 3 years | 14 | 7.6 |
|  | > 3 years | 7 | 3.8 |

In relation to the coping strategies used, higher means were perceived in relation to general stress rather than stress related to gender identity (Table 2). Considering the total score of the coping scale (sum), there is no significant relationship with quality of life, both for general stress and for stress related to gender identity.

**Table 2.** Mean (SD) coping general stress and gender-related stress.

|  |  | Mean (SD) |
| --- | --- | --- |
| **Coping general stress** | Hobby<sup>a</sup> | 3.09 (0.09) |
|  | Socialization<sup>b</sup> | 3.08 (0.13) |
|  | Introspection<sup>c</sup> | 2.80 (0.12) |
|  | Avoidance<sup>d</sup> | 3.29 (0.35) |
| **Coping gender-related stress** | Hobby<sup>a</sup> | 1.22 (0.11) |
|  | Socialization<sup>b</sup> | 1.61 (0.12) |
|  | Introspection<sup>c</sup> | 1.42 (0.11) |
|  | Avoidance<sup>d</sup> | 1.76 (0.13) |

<sup>a</sup>Total of 5 strategies.
<sup>b</sup>Total of 8 strategies.
<sup>c</sup>Total of 7 strategies.
<sup>d</sup>Total of 6 strategies.

Regarding quality of life, 53.9% of the sample was below the median, representing low QoL (Table 3). As for the mental health variables, the sample showed 48.0% with significant presence of depressive symptoms, 68.3% had significant symptoms of anxiety and 49.6% had some degree of gender dysphoria (GD) – “with GD” and “with a lot of GD”.

**Table 3.**
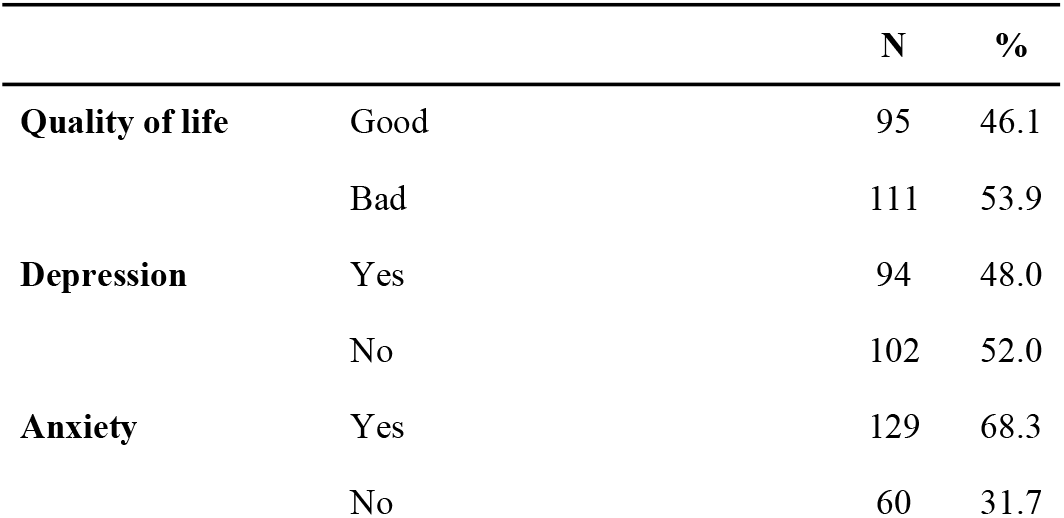
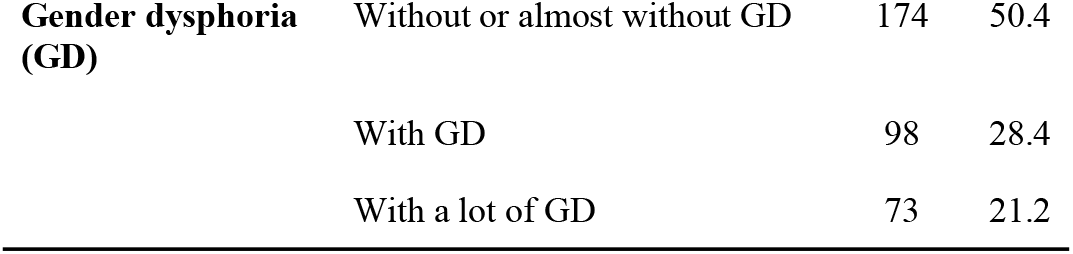
Mental health variables.

|  |  | N | % |
| --- | --- | --- | --- |
| **Quality of life** | Good | 95 | 46.1 |
|  | Bad | 111 | 53.9 |
| **Depression** | Yes | 94 | 48.0 |
|  | No | 102 | 52.0 |
| **Anxiety** | Yes | 129 | 68.3 |
|  | No | 60 | 31.7 |
| **Gender dysphoria (GD)** | Without or almost without GD | 174 | 50.4 |
|  | With GD | 98 | 28.4 |
|  | With a lot of GD | 73 | 21.2 |

When grouped together, avoidance strategies were associated with a worse quality of life, in relation to general stress; whereas the other dimensions (hobby, socialization, and introspection) were associated with a better quality of life (Table 4). There was no significant association between coping strategies for stress related to gender identity and quality of life.

**Table 4.** Correlation quality of life and coping general stress and gender-related stress.

|  |  | **p** |
| --- | --- | --- |
| **Coping general stress** | Hobby | 0.036<sup>a</sup> |
|  | Socialization | 0.170<sup>b</sup> |
|  | Introspection | 0.177<sup>b</sup> |
|  | Avoidance | -0.194<sup>a</sup> |
| **Coping gender-related stress** | Hobby | 0.015 |
|  | Socialization | -0.001 |
|  | Introspection | 0.043 |
|  | Avoidance | -0.102 |

<sup>a</sup>p<0.001.
<sup>b</sup>p<0.05.

Two Poisson regressions were performed in two steps with the quality of life outcome. The first analysis concerned coping strategies with general stress and the second, with coping strategies for stress related to gender identity (Table 5). In the first step, only the four dimensions of coping strategies were used. In the second step, for both analyses, the variables for depressive symptoms, anxiety symptoms, gender dysphoria, age, and socioeconomic status were included.

**Table 5.**
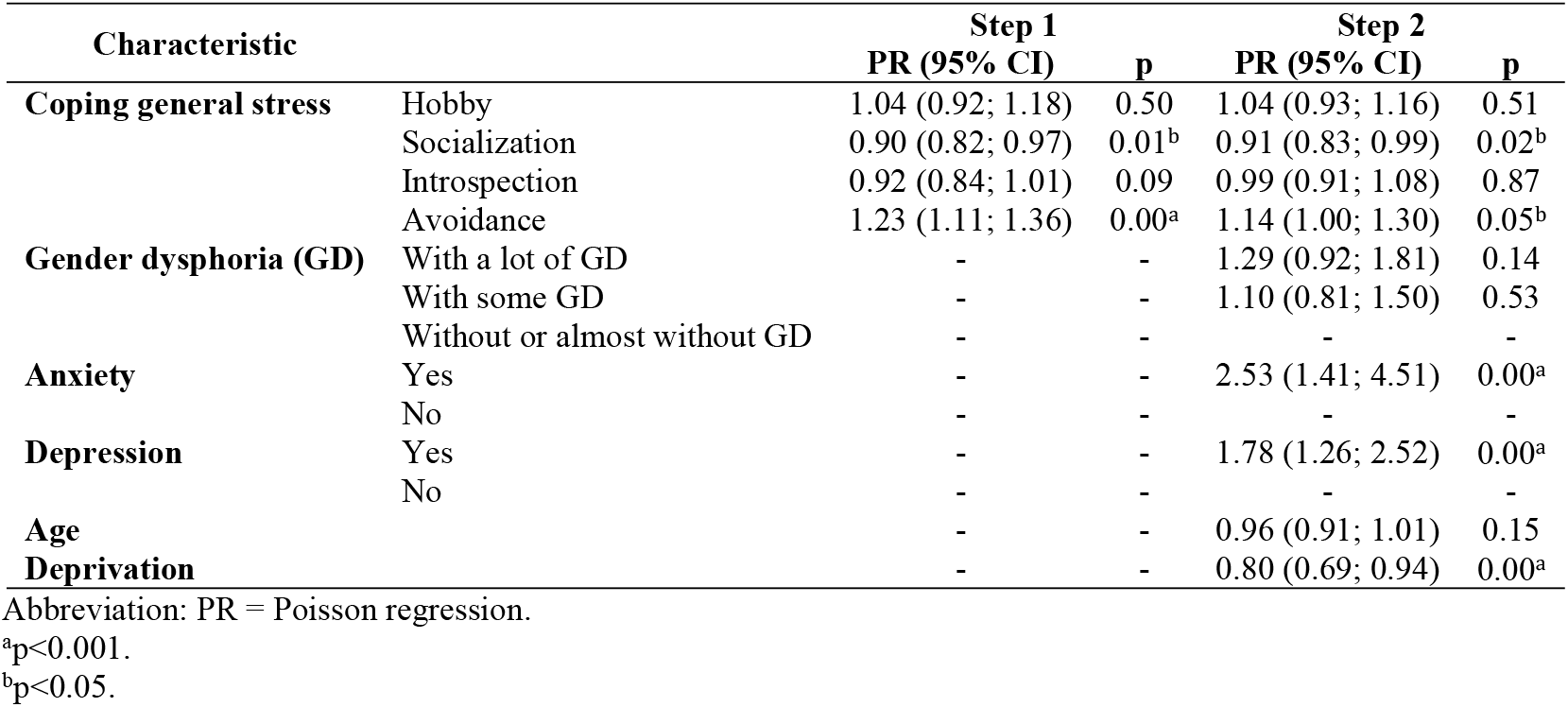
Poisson regression quality of life and coping general stress.

| Characteristic |  | Step 1 |  | Step 2 |  |
| --- | --- | --- | --- | --- | --- |
|  |  | PR (95% CI) | p | PR (95% CI) | p |
| **Coping general stress** | Hobby | 1.04 (0.92; 1.18) | 0.50 | 1.04 (0.93; 1.16) | 0.51 |
|  | Socialization | 0.90 (0.82; 0.97) | 0.01<sup>b</sup> | 0.91 (0.83; 0.99) | 0.02<sup>b</sup> |
|  | Introspection | 0.92 (0.84; 1.01) | 0.09 | 0.99 (0.91; 1.08) | 0.87 |
|  | Avoidance | 1.23 (1.11; 1.36) | 0.00<sup>a</sup> | 1.14 (1.00; 1.30) | 0.05<sup>b</sup> |
| **Gender dysphoria (GD)** | With a lot of GD | - | - | 1.29 (0.92; 1.81) | 0.14 |
|  | With some GD | - | - | 1.10 (0.81; 1.50) | 0.53 |
|  | Without or almost without GD | - | - | - | - |
| **Anxiety** | Yes | - | - | 2.53 (1.41; 4.51) | 0.00<sup>a</sup> |
|  | No | - | - | - | - |
| **Depression** | Yes | - | - | 1.78 (1.26; 2.52) | 0.00<sup>a</sup> |
|  | No | - | - | - | - |
| **Age** |  | - | - | 0.96 (0.91; 1.01) | 0.15 |
| **Deprivation** |  | - | - | 0.80 (0.69; 0.94) | 0.00<sup>a</sup> |

Abbreviation: PR = Poisson regression.
<sup>a</sup>p<0.001.
<sup>b</sup>p<0.05.

In the first Poisson regression, a worse quality of life was related to avoidance strategies in dealing with general stress, whereas socialization coping strategies were associated with a better quality of life. Both relationships were maintained when the other predictor variables were added to the analysis. In the second step, low socioeconomic status and the presence of anxiety and depression symptoms were significantly associated with a worse quality of life.

In the second Poisson regression, there was a significant association between a worse quality of life and avoidance strategies for coping with gender-related stress (Table 6). This relationship was not statistically significant when the other predictor variables were added. In the second step, low socioeconomic status and the presence of anxiety and depression symptoms were significantly associated with a poorer quality of life.

**Table 6.** Poisson Regression quality of life and coping gender-related stress.

| Characteristic |  | Step 1 |  | Step 2 |  |
| --- | --- | --- | --- | --- | --- |
|  |  | PR (95% CI) | p | PR (95% CI) | p |
| **Coping gender-related stress** | Hobby | 0.92 (0.81; 1.05) | 0.23 | 1.00 (0.90; 1.11) | 0.98 |
|  | Socialization | 0.99 (0.89; 1.10) | 0.88 | 1.01 (0.92; 1.11) | 0.87 |
|  | Introspection | 0.95 (0.85; 1.07) | 0.38 | 0.95 (0.85; 1.07) | 0.44 |
|  | Avoidance | 1.15 (1.03; 1.30) | 0.01<sup>b</sup> | 1.01 (0.91; 1.12) | 0.84 |
| **Gender dysphoria** | With a lot of GD | - | - | 1.27 (0.91; 1.79) | 0.16 |
|  | With some GD | - | - | 1.13 (0.84; 1.53) | 0.42 |
|  | Without or almost without GD | - | - | - | - |
| **Anxiety** | Yes | - | - | 2.56 (1.38; 4.74) | 0.00<sup>a</sup> |
|  | No | - | - | - | - |
| **Depression** | Yes | - | - | 1.85 (1.27; 2.68) | 0.00<sup>a</sup> |
|  | No | - | - | - | - |
| **Age** |  | - | - | 0.97 (0.91; 1.03) | 0.27 |
| **Deprivation** |  | - | - | 0.79 (0.68; 0.93) | 0.00<sup>a</sup> |

Abbreviation: PR = Poisson regression.
<sup>a</sup>p<0.001.
<sup>b</sup>p<0.05.

## Discussion

This study highlights the challenges that Brazilian transgender youth face when dealing with the stress of everyday life and of social prejudice due to their gender status. Our study confirmed that more than half of the sample showed a self-perceived low quality of life (53.9%). Other studies have shown similar results (Jones et al., 2019; Newfield et al., 2006; Nobili et al., 2018). Despite being a subjective concept that involves different social and individual values, this self-perceived low quality of life raises questions concerning the main challenges that transgender youth face and what we must do, as a society, to minimize their suffering. Despite the social advances of recent years regarding sexuality, identifying as transgender is still a stressful and dysfunctional condition.

In accordance with other studies (Budge, Adelson, & Howard, 2013; Lewis et al, 2019; Müller et al., 2019), we found high rates of depressive symptoms (48.0%) and anxiety symptoms (68.3%). As predicted, these mental conditions were significantly associated with poor self-perceived quality of life, in addition to coping with general stress and gender-related stress. Generally, the Brazilian population shows a higher prevalence of depression (5.8%) and anxiety (9.3%) disorders (WHO, 2017) among South Americans countries and a probable lower quality of life; the worst prevalence of mental conditions, however, occurs within the transgender youth population. The Brazilian study with trans and gender non-conforming (TGNC) adults found high rates of depressive symptoms (67.2%), indicating that suffering may persist throughout adult life (Chinazzo et al., 2021). Another predictive variable associated with low quality of life in both regression analysis was deprivation, that may harm any population enduring difficult social conditions. Socioeconomical factors are associated with mental health and low quality of life, especially in a stigmatized population.

Quality of life from a personal perspective can be improved with appropriate coping strategies to buffer the general stressors of life. Coping strategies can be learned and developed and the concept of adaptive and maladaptive strategies depends on the impact on well-being and mental health of each individual (Lazarus & Folkman, 1984). Transgender people may need more specific coping strategies. A central aspect for our research was to look at strategies that may be adaptive or not for Brazilian transgender youth. In this study, the coping strategies were grouped into four clusters: hobby, socialization, introspection, and avoidance and two types of stress: general and gender-related. These two types of stress were analyzed to understand specific transgender stressors in relation to quality of life in comparison with the general stresses of life that they also experience, and to know which adaptive coping strategies may buffer the dysfunctional consequences of adverse stressful events.

Our final findings concerning coping strategies for general stress indicated a significant association between a lower quality of life with less use of socialization (p=0.02) and more use of avoidance (p=0.05). Socialization coping strategies such as passing time with family, friends, and volunteer work was associated with better self-perception and quality of life, which may indicate that these social relations are positive and protective for transgender youth. Nevertheless, this variable was not related to public health services support.

In another study published by our group (Chinazzo et al., 2021), we found that there was an association between supportive affective relationships with a diminished presence of depressive symptoms and suicidal ideation in transgender Brazilian adults. Despite the social difficulties related to prejudice faced by transgender individuals, in our sample we identified socialization as an adaptative coping strategy for general stress. This finding highlights the need for a special social resource and strategies to reduce the chances of serious mental health issues in transgender youth.

Avoidance coping strategies for general stress were related with a low self-perceived quality of life. Despite the possible passing pleasure associated with these coping strategies (excessive sleeping, eating, shopping, and superfluous self-care), these methods of coping draw them away from a more mature solution to their challenges in dealing with general stress and as a result, compromise their mental health. This result implies that clinics working with transgender should work on diminishing these maladaptive strategies. The use of maladaptive coping strategies has been previously reported in transgender populations and is implicated in contributing to serious mental health issues (Freese et al., 2017; Hughto et al., 2017).

Surprisingly, our study did not find a significant association between quality of life and the four clusters of coping mechanisms to deal with gender-related stress. In the first step of the regression analysis, the use of avoidance coping strategies was significantly associated with a low perceived quality of life (p=0.01). However, this association is not apparent in the second step, in which other variables were also included.

Avoidance strategy is associated with social isolation, marginalization, and failure to find personal solutions; as cited before, this strategy may lead to anxiety, depression, suicidal behavior, and, as evidenced by our study, poor perceived quality of life. The Brazilian social environment is still full of different expressions of prejudice and aggressive attitudes directed towards transgender people. It is rational to think that avoidance coping strategies could be considered as “protective” from some points of view, as suggested by the Minority Stress Theory. It is a challenge for medical assistance teams to help change this “safe” coping strategy for a more “audacious one.” For these changes in coping attitudes to occur, the transgender youth will need to develop emotional strength and self-assurance, aspects uncommon in many youths.

In our study we found a low average use of coping mechanisms when the four clusters and gender-related stress were considered. This low average will need to be studied in the future to better understand the coping mechanisms used by transgender youth in Brazil. Also, a further study should explore the relation between quality of life, coping, and gender-related stress in the transgender population.

### Limitations

To the best of our knowledge, this is the first study with Brazilian transgender youth from different regions of the country, which includes quality of life and coping strategies for general and gender-related stress. There were some statistical limitations in generalizing the results found in the Brazilian binary and non-binary transgender youth population due to the relatively small sample studied. Nevertheless, the on-line survey gave access to many regions of Brazil which allowed for a heterogeneous sample, albeit a small one.

Our study included binary and non-binary transgender youth. However, the sample was not enough to calculate the difference between the two groups, binary and non-binary. Although both groups are included under the transgender umbrella, there is evidence that there may exist different stressors, such as quality of life and mental health issues (Jones, et al., 2019). To better support both groups, we suggest more studies in this area.

## Data Availability

The database includes hospital patient data, for ethical reasons, the data is available via email with the authors.

## Acknowledgments

We thank the Health Equity and Epidemiology Research Group (HEER) for multiple, and very useful, insights. We also appreciate TransYouth CAN! Team for sharing their survey. National Council for Scientific and Technological Development (CNPq) and Coordination for the Improvement of Higher Education Personnel (CAPES) provided financial support through scholarships. The authors thank the Research Incentive Fund of the Hospital da Clínicas de Porto Alegre (FIPE/HCPA) for the financial support.

